# Development and validation of direct RT-LAMP for SARS-CoV-2

**DOI:** 10.1101/2020.04.29.20075747

**Authors:** Abu Naser Mohon, Jana Hundt, Guido van Marle, Kanti Pabbaraju, Byron Berenger, Thomas Griener, Luiz Lisboa, Deirdre Church, Markus Czub, Alexander Greninger, Keith Jerome, Cody Doolan, Dylan R. Pillai

## Abstract

We have developed a reverse-transcriptase loop mediated amplification (RT-LAMP) method targeting genes encoding the Spike (S) protein and RNA-dependent RNA polymerase (RdRP) of SARS-CoV-2. The LAMP assay achieves comparable limit of detection as commonly used RT-PCR protocols based on artificial targets, recombinant Sindbis virus, and clinical samples. Clinical validation of single-target (S gene) LAMP (N=120) showed a positive percent agreement (PPA) of 41/42 (97.62%) and negative percent agreement (NPA) of 77/78 (98.72%) compared to reference RT-PCR. Dual-target RT-LAMP (S and RdRP gene) achieved a PPA of 44/48 (91.97%) and NPA 72/72 (100%) when including discrepant samples. The assay can be performed without a formal extraction procedure, with lyophilized reagents which do need cold chain, and is amenable to point-of-care application with visual detection.

## Introduction

Over the last several decades, we have witnessed the rise of both known and novel viruses, including human immunodeficiency virus (HIV), SARS coronavirus (SARS-CoV), MERS-CoV, influenza H1N1, Ebola virus (EBOV), Dengue (DENV), Chikungunya (CHIK), Zika (ZIKV), and most recently 2019 novel coronavirus (SARS-CoV-2/COVID-19)(1). Most of these emerging viral infections have been triggered by a direct zoonotic (animal-to-human) transmission event or enhancement, proliferation and spread of vectors such as the mosquito in new geographic areas. In December 2019 and early January 2020, a cluster of pneumonia cases from a novel coronavirus, SARS-CoV-2, was reported in Wuhan, China (2–4).

SARS-CoV-2 has now resulted in a global pandemic with the epicentre at the time of writing in Europe and North America (5). A common theme in the public health response to COVID19 and similar threats is the lack of rapidly deployable testing in the field to screen large numbers of individuals in exposed areas, international ports of entry, and testing in quarantine locations such as the home residences, as well as low-resourced areas (6). This hampers case finding and increases the number of individuals at risk of exposure and infection. With the ease of travel across continents, delayed testing and lack of screening programs in the field, global human-to-human transmission will continue at high rates. These factors make a pandemic very difficult to contain. Early identification of the virus and rapid deployment of a targeted point of care test (POCT) can stem the spread through immediate quarantine of infected persons(7). We used existing viral genome sequences to develop a SARS-CoV-2 loop mediated amplification (LAMP) assay for clinical use and evaluated whether an extraction-free and instrument-free approach could be achieved(8, 9). POCT requires portability and low complexity without reliance on sophisticated extraction and read-out instrumentation. Furthermore, LAMP relies on an alternate set of reagent chemistry that does not depend on or hinder critical elements of the RT-PCR supply chain which is now under duress (10). Our group has previously demonstrated the utility of LAMP for other infectious agents like malaria and dengue (11–13).

## Materials and Methods

### Patient samples and Ethics

Clinical samples used in this study were standard nasopharyngeal (NP) swabs in viral transport medium (VTM). Specifically, archival samples were sourced from the University of Washington, Seattle, USA and extracted viral RNA from the Alberta Public Health Laboratory. Ethical approval for use of the archived samples was obtained from the Conjoint Health Research Ethics Board (CHREB) of the University of Calgary (REB20–0402). The use of de-identified specimens was deemed non-human subject work by the University of Washington Institutional Review Board (IRB).

### LAMP primer design

Genomic sequences (cDNA) of the SARS-CoV-2 were retrieved from the GenBank database (https://www.ncbi.nlm.nih.gov/genbank/sars-cov-2-seqs/) and multiple sequence alignment analysis (https://www.ebi.ac.uk/Tools/msa/clustalo/) was conducted with other related viruses. From the multiple sequence alignment, several regions unique to the SARS CoV-2 were identified using our own algorithms developed in collaboration with Illucidx Inc. (Calgary, AB). LAMP primer sets were designed targeting unique regions of the Spike (S) protein gene, RNA-dependent RNA Polymerase gene (RdRP), and parts of the Open Reading Frame (ORF) 1a/b. Five sets of LAMP primers were selected for laboratory analysis, where Set 1 and Set 2 targeted the non-structural protein (nsp) 3 region in the ORF1a/b gene and S protein gene, respectively; and Sets 3, 4, and 5 were designed to amplify different regions of RdRP gene of SARS CoV-2 (Table 1). For the external LAMP amplification control, primers were used against bacteriophage MS2 as previously described (14).

**Table 1:**
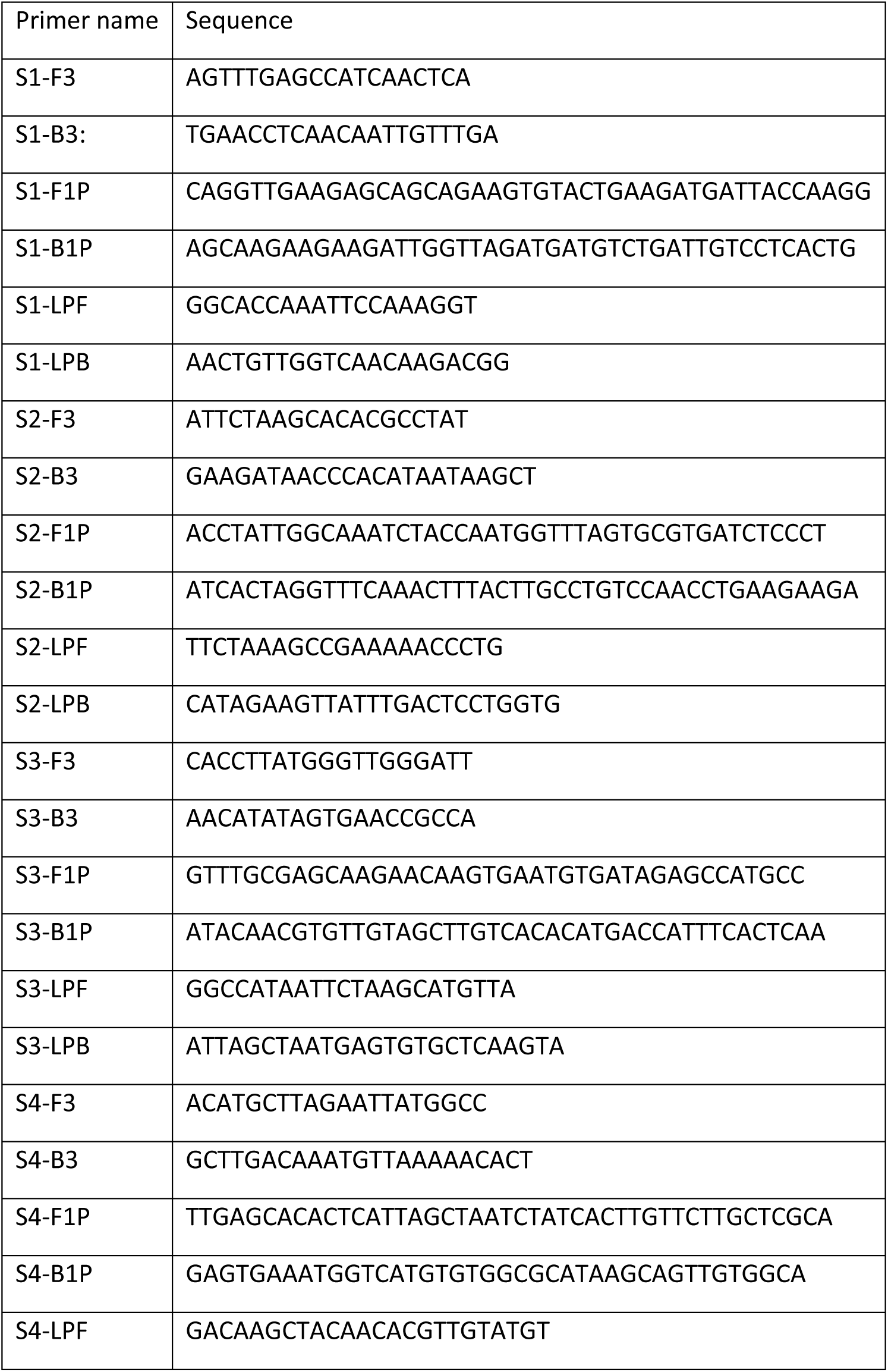
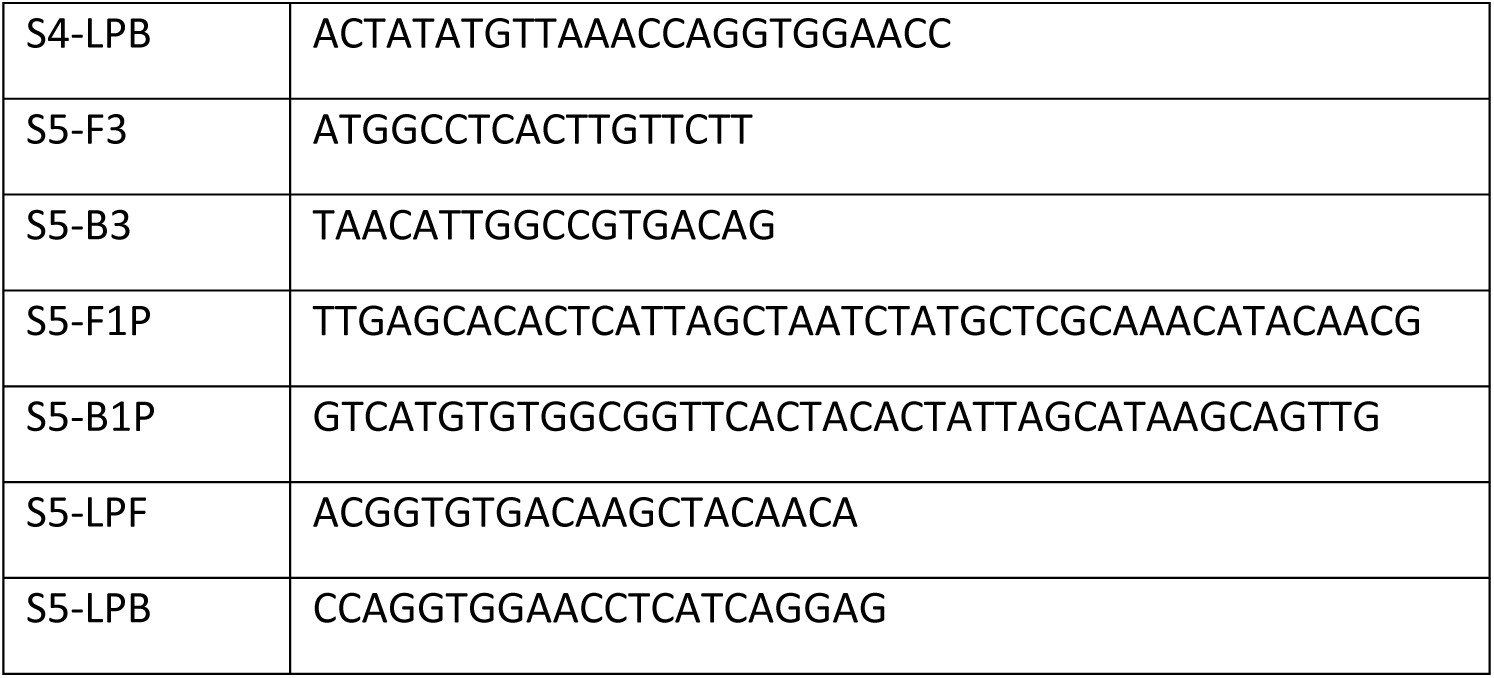
Primer sets used in this study to perform RT-LAMP. All 5 primer sets are shown: Set 1 (ORF1a/b, nsp3); Set 2 (S gene); and Set 3, 4, and 5 (RdRP).

### Design of artificial viral target for LAMP assay verification

Four fragments of specific SARS-Co-V2 regions (ORF1ab (nsp 3,10–11), RdRP (nsp 12), and spike (S)) were synthesized by SGI-DNA Inc. (San Diego, CA). Fragments were ligated to make one large concatenated DNA template using the BioXP3200 (SGI-DNA, San Diego, CA) automated Gibson assembly system. The final template was 1097 base pairs long containing concatenated ORF1a/b (nsp 3)-Spike protein-RdRP (nsp 12) - ORF1a/b (nsp 10–11) fragments together with flanking plasmid sequence in that order.

Reverse transcription PCR (RT-PCR) assays used in this study

The RT-PCR assays used in this study were performed according to previous publications for the E gene(15) and N2 gene(16). The E gene RT-PCR was performed with modification according to the Alberta Public Health Laboratory reference method (17). The modifications included the addition of GC clamps at the 3’ end of the primers and the shortening and addition of a minor groove binding (MGB) moiety to the hydrolysis probe. Five (5) μL of input template was used to perform the RT-PCR reactions on the same day when RT-LAMP was performed. A maximal Ct value cut-off of 40 was used to determine positivity for all RT-PCR reactions.

### LAMP assay conditions

The single-target LAMP reaction was conducted using a combination of Warmstart Rtx Reverse Transcriptase (New England Biolab, Whitby, ON) with *Bst 2.0* Warmstart DNA Polymerase (New England Biolab, Whitby, ON). In a 25 µL LAMP reaction mixture, 1.6 µM F1P and B1P, 0.8 µM LPF and LPB, 0.2 µM F3 and B3 primer concentrations, 8mM MgSO_4_, 1.4 mM dNTPs, 8 unit of *Bst* 2.0 WarmStart^®^ DNA Polymerase and 7.5 unit of Warm Start® reverse transcriptase were used. The assay was optimized with pre-addition of 0.5µL of 50X SYBR green (Invitrogen, Burlington, ON) in the 25 µL reaction mixture. For all LAMP experiments, 10 µL of template was used in the 25 µL reaction mixture. For dual-target LAMP, identical reagent composition was used except the primers (6 per target) that were added at double concentration and one half the volume for each primer set. Amplification was measured through increased relative fluorescence units (RFU) per minute in the CFX-96 Real-Time PCR detection system (Bio-Rad, Mississauga, ON) or fluorescence values on the QuantStudio 5 RT system (ThermoFisher, Toronto, ON). RFU values above 100 on the CFX-96 instrument or greater than 100,000 on the QuantStudio 5 instrument were considered positive when associated with a typical amplification curve before a 30-minute reaction time. Each primer set was used to amplify the artificial viral DNA target at different temperature (61°C, 63°C, and 65°C) for a maximum LAMP assay run time of 40 minutes.

### Visual detection of the LAMP without instrumentation

A positive LAMP assay was detected visually by the pre-addition of 0.5µL colorimetric fluorescence indicator (CFI). CFI was made up of the combination of 0.7% (v/v) 10000X Gelgreen (Biotium, Freemont, CA) in 12 mM Hydroxynapthol blue resuspended in dH_2_0 (Sigma-Aldrich, Oakville, ON). After the 30-minute LAMP reaction time, the tubes were exposed to blue LED light using a Blue Light Transilluminator (New England Biogroup, Atkinson, NH) to visualize the green fluorescence.

### Lyophilized LAMP without the need for cold chain

In order to determine if the LAMP primers and master mix could be lyophilized, oligonucleotides were shipped to Pro-Lab Diagnostics (Toronto, Canada) and lyophilized with GspSSD2 isothermal master mixture (Optigene, UK). The lyophilized primer, enzyme, master mix combination was hydrated in 15 μL of resuspension buffer (Pro-Lab Diagnostics) to which 10 μL of the sample was added. Both direct LAMP (see method described later) and kit-based RNA extractions were performed in this way.

### Limit of detection studies

Limit of detection of the LAMP assay was evaluated in three different ways. Initially, copy number of the synthesized DNA fragment was determined by comparing concentration and molecular weight.

Subsequently, the template solution was serially diluted to achieve a range from 5,000,000 to 5 copies per reaction. These serially diluted templates were tested by all primers sets. Secondly, the extracted RNA from one patient specimen was 10-fold serially diluted and tested with the LAMP assay and two RTPCR assays used by reference laboratories targeting both the E gene(15) and N2 gene(16). Finally, the artificial template containing the targeted sequences of interest was cloned into Sindbis Virus (SV) viral vector system (SINrep5) containing green fluorescent protein (EGFP) and then transfected into BHK-21 cell lines(18),(19). An approximate estimation of the viral titer was determined by comparing EGFP expressing foci forming units in BHK-21 as described previously. EGFP forming units ranging from 10^6^ – 10^8^/mL were obtained for various recombinant virus stocks. Maintenance of the SARS/CoV2-targetted sequences in the recombinant virus was confirmed using RT-PCR with primers targeting the flanking SV sequences. Virus particles were serially diluted from 100 to 0.001 SV EGFP forming units per μL, and RNA was extracted with the QiaAmp Viral RNA extraction kit (Qiagen, Toronto, ON). Extracted RNA was subjected to TURBO™ DNase (ThermoFisher, Toronto, ON) digestion. Dilutions were subjected to LAMP reactions as described above.

### Validation using clinical samples

In total, 42 RT-PCR-positive clinical (n=32), contrived (n=10), and 78 negative NP swab samples were tested in the single-target S gene LAMP validation study. In the negative panel, four common human coronavirus RNA (strain 0C43, NL63, 229E, and HKU1), respiratory syncytial virus (RSV), and Influenza H1N1 clinical samples were included to evaluate the specificity of the test. Contrived samples were generated by inoculating the artificial gene DNA construct described earlier into VTM from NP swabs. Extraction of RNA from clinical samples was performed using the NUCLISENS easyMAG system (Biomerieux, Durham, NC) or QIAamp Viral RNA Mini Kit (Qiagen, Toronto, ON) depending on specimen source. All samples in the set were tested simultaneously using the E gene RT-PCR described above and S gene LAMP (primer set 2). Discrepant analysis was performed by performing the CDC N2 gene(16) RT-PCR assay. For the LAMP assay, 10 μL of the RNA extract was used for each reaction. A second validation study was performed for the dual-target LAMP (S gene and RdRP). Here, RT-PCR-positive clinical (n=34), contrived (n=10), and 72 negative NP swab samples were tested. All LAMP reactions for the clinical validation studies was performed using the combination of Warmstart Rtx Reverse Transcriptase with Bst 2.0 Warmstart DNA Polymerase as described earlier.

### Direct LAMP assay without formal extraction

A direct LAMP assay was also conducted to establish an extraction-free approach. In this scheme, the LAMP reaction mixture was prepared without the enzymes. 9.5 µL reaction mixture containing all reagents except the enzymes was added to 14 µL of the 10-fold serially diluted NP VTM sample. For direct LAMP, VTM must be diluted 1:10 (v/v) with dH_2_0 prior to addition. The mixture was then heated at 95°C for 3, 5, and 10 minutes to both inactivate virus and presumptively release viral RNA. Finally, *Bst* 2.0 WarmStart^®^ DNA Polymerase (1 µL) and Warm Start® reverse transcriptase (0.5 µL) were directly added to the reaction mixture and the LAMP assay was carried out as above. Due to evaporative loss, boil steps should have excess volume to ensure adequate input template for LAMP reactions.

### In silico analysis of primer combinations to determine cross-reactivity

A blast search alignment (https://blast.ncbi.nlm.nih.gov/Blast.cgi) for primers in set 2 (spike gene) and set 3 (RdRP gene) were performed against a critical list of infectious agents that cause upper respiratory tract infections. A nucleotide local alignment using BLASTn with the default parameters was performed against the National Center of Biotechnology Information (NCBI) Nucleotide database.

## Results

### Verification of SARS-CoV-2 LAMP primer sets on artificial gene targets

In order to determine the limits of detection of the five LAMP primer sets designed for SARS-CoV-2, experiments were conducted using an artificial gene target construct. Three targets (Figure 1) selected were the Spike (S) protein gene, the RNA-dependent RNA polymerase (RdRP) region (nsp 12), and nsp 3 region in the open reading frame (ORF) 1a/b protein encoding gene sequence. Figure 2 shows the amplification curves for the five primer sets used in this study. Based on time to positivity, primer sets 1, 2, and 3 demonstrated the fastest positive reaction time, suggesting optimal performance in the LAMP assay at 50 copies of the artificial target per reaction. When tested at 5 copies per reaction, primer set 2 targeting the S gene showed the best limit of detection. An *in silico* analysis of primer set 2 (S gene) and set 3 (RdRP) demonstrated no significant sequence alignment cross-reactivity with known upper respiratory tract infectious pathogens (data not shown).

**Figure 1:**
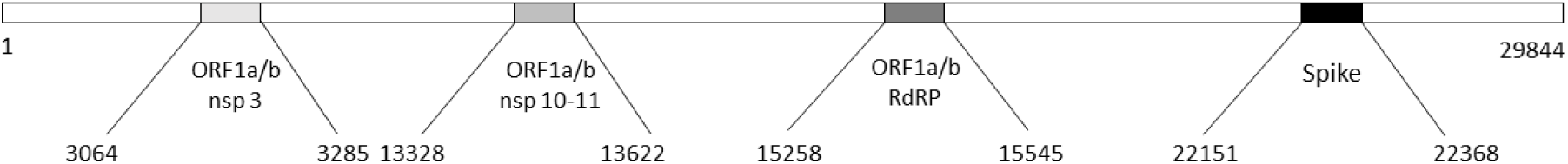
Map of the gene fragments from SARS-Co-V2 (Genbank ID MT2078.1) that were used for synthesizing the genetic construct template. Fragments of ORF1a/b [nsp 3] (3064–3285), ORF1a/b [nsp 10–11] (13328–13622), ORF1a/b RdRP [nsp 12] (15258–15445), and Spike gene (22153–22368) were concatenated into a single artificial construct. A second template including the E gene was also made for the purposes of testing the E gene RT-PCR.

**Figure 2:**
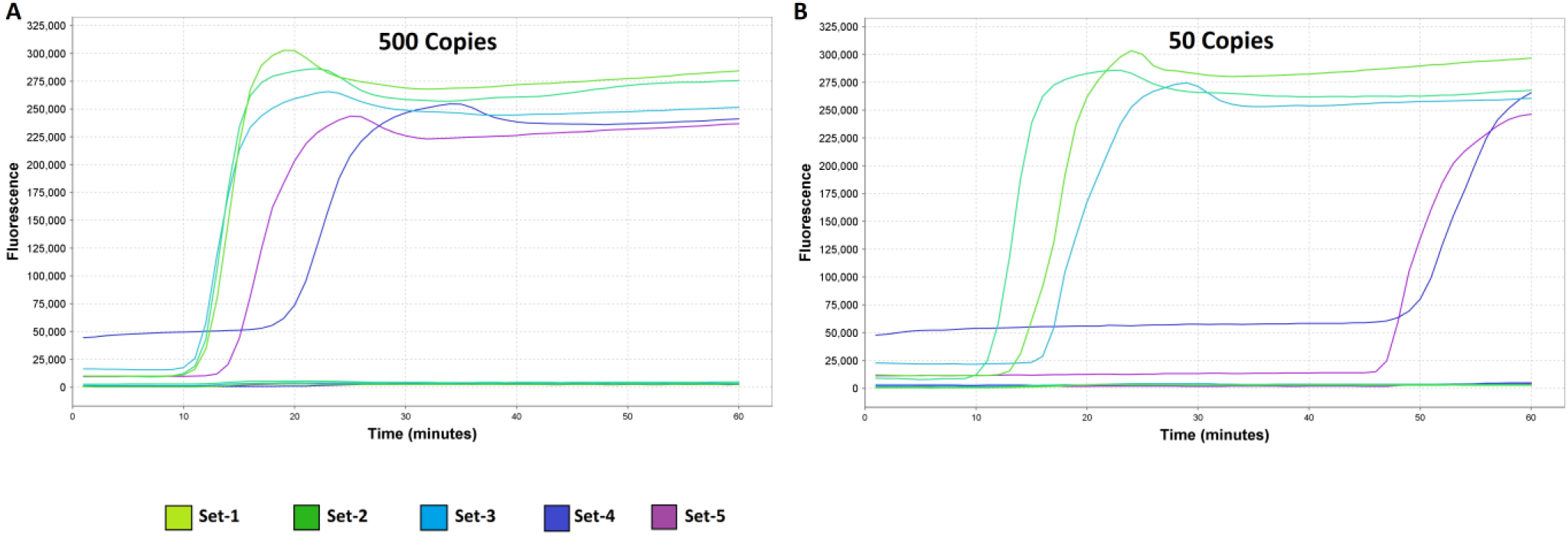
Limit of detection of RT-LAMP primer sets designed to detect the SARS-CoV-2 artificial DNA construct. These data are representative of an experiment performed in triplicate at (A) 500 and (B) 50 copies. Amplification curves for all 5 primer sets are shown: Set 1 (ORF1ab); Set 2 (S gene); and Set 3, 4, and 5 (RdRP). Based on these experiments, a cut off 30 minutes reaction time was determined for specific amplification. Fluorescence (QuantStudio 5) on the y-axis is plotted in relation to reaction time (minutes).

### Visual detection of SARS-CoV-2 LAMP amplification

An advantage of LAMP is the ability to detect amplification with the naked eye either via the use of colorimetric or fluorescent dyes. To test this, LAMP was conducted using the S gene LAMP primer set using the artificial gene target. Figure 3 shows a positive test based on fluorescent detection using a blue light emitting diode (LED). A serial dilution between 50 × 10^7^ and 5 copies per reaction of the artificial gene construct is shown with a limit of detection of 50 copies per reaction obtained. The LAMP assay using enzyme GspSSD2 was also performed using a lyophilized master mix. Studies were performed with both kit-based RNA extracted clinical samples as well as using the direct LAMP method described later. These data showed that amplification occurred up to a 1000-fold dilution of a clinical sample with the direct LAMP method (data not shown).

**Figure 3:**
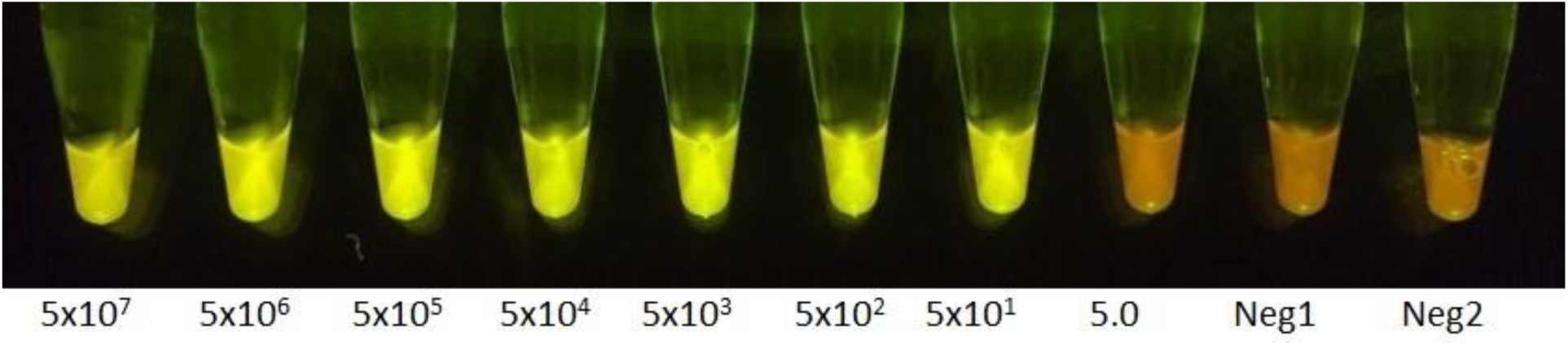
Photograph of S gene RT-LAMP (Set 2) performed on the artificial DNA construct. Fluorescence can be detected by naked eye after excitation of gel green in the reaction with a blue LED light. A serial 10-fold dilution is shown from 5 × 10^7^ copies to 5 copies of the gene construct in this representative experiment (left to right). The last two tubes on the right are negative template controls. A reaction time was set at 30 minutes.

### Limit of detection studies using recombinant Sindbis virus (SV) containing SARS-CoV-2 targets

In order to determine LOD based on viral titer, recombinant RNA viral vector from Sindbis virus (SV) containing SARS-CoV-2 targets was generated. LAMP was performed using Set 2 (S gene) alone and in combination with Set 3 (RdRP region (nsp 12)). The primer sets in question achieved a LOD of 0.1–1.0 SV EGFP forming units per μL for the S gene alone and the dual-target LAMP amplifying both S and RdRP genes in a single reaction (Table 2). One SV EGFP focus forming unit (FFU) roughly corresponds to 1.0 infectious viral particle given the assumption that one virus particle infects one cell.

**Table 2:** RT-LAMP assay results on serially diluted recombinant SV virus containing the artificial construct now expressed as RNA. Positive tested replicates/total number of replicates is shown in relation to the approximate viral titer for both single-target (S gene) and dual-target (S and RdRP gene) LAMP reactions.

| Approximate viral titer/μL | S gene<br>Single-target RT-LAMP | S gene and RdRP gene<br>Dual-Target RT-LAMP |
| --- | --- | --- |
| 100 | 3/3 | 3/3 |
| 10 | 3/3 | 3/3 |
| 1 | 3/3 | 3/3 |
| 0.1 | 2/3 | 2/3 |
| 0.01 | 0/3 | 1/3 |
| 0.001 | 0/3 | 0/3 |

### Direct LAMP detection of SARS-CoV-2 without formal extraction

Given extraction reagent supply chain shortages, we tested the ability of LAMP using the S gene to amplify a SARS-CoV-2 positive nasopharyngeal swab specimen without a formal extraction procedure. The VTM was heated at 95°C for 3, 5 and 10 minutes and then subjected to the S gene LAMP procedure in a serial dilution experiment. This experiment was performed in a head-to-head comparison with the same specimen tested after formal extraction. Figure 4 demonstrates that amplification occurred with direct LAMP up to a dilution of 100,000-fold with a 95°C for 3 minutes heat step in a single experiment. Table 3 shows the data for direct LAMP compared to LAMP and RT-PCR from RNA extracts in triplicate experiments. Reproducible amplification with direct LAMP (95°C for 3 minutes) occurred at a dilution factor of 1,000-fold, whereas LAMP from an RNA extract was successful reproducibly at a dilution of 100,000-fold.

**Figure 4:**
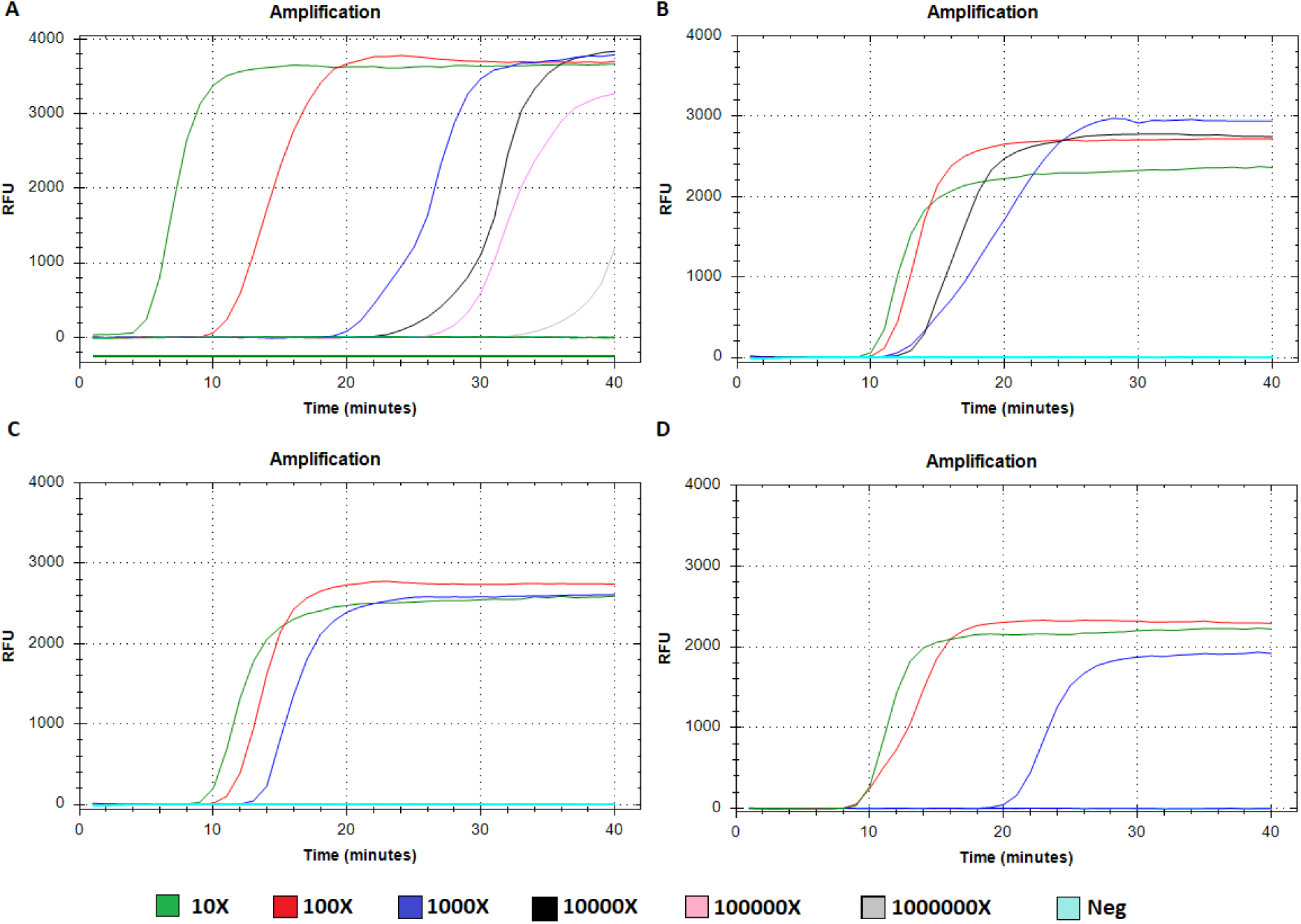
Single-target (S gene) RT-LAMP amplification of SARS-CoV-2 spike gene from NP swab sample (E gene Ct 21.29) using a simple heat step. A representative direct LAMP experiment is compared to a kit-based extracted RNA (A) for serial dilutions of the neat sample. Heat inactivation without formal extraction is shown in (B) 95°C for 3 minutes (C) 95°C for 5 minutes and (D) 95°C for 10 minutes. Relative fluorescence units (CFX-96) on the y-axis is plotted in relation to reaction time (minutes).

**Table 3:** Single-target (S gene) RT-LAMP assay results on a NP swab VTM sample (E gene Ct 21.29) containing SARS-Co-V2 compared to RT-LAMP and RT-PCR after kit-based RNA extraction. Positive tested replicates/total number of replicates for a single representative triplicate experiment is shown. Dilutions are based on 10-fold serial dilutions from a neat NP VTM sample. Kit-based extraction is compared to direct RT-LAMP procedures including a heat step.

| Sample dilution factor | E gene<br>RNA extract<br>RT-PCR | N2 gene<br>RNA extract<br>RT-PCR | RT-LAMP<br>RNA extract | Direct RT-LAMP<br>10 min/95°C | Direct RT-LAMP<br>5 min/95°C | Direct RT-LAMP<br>3 min/ 95°C |
| --- | --- | --- | --- | --- | --- | --- |
| 10 | 3/3 | 3/3 | 3/3 | 3/3 | 3/3 | 3/3 |
| 100 | 3/3 | 3/3 | 3/3 | 3/3 | 3/3 | 3/3 |
| 1000 | 3/3 | 3/3 | 2/3 | 1/3 | 2/3 | 2/3 |
| 10000 | 3/3 | 3/3 | 3/3 | 0/3 | 0/3 | 1/3 |
| 100000 | 2/3 | 2/3 | 2/3 | 0/3 | 0/3 | 1/3 |

### Limit of detection studies using SARS-CoV-2 LAMP on nasopharyngeal swab clinical samples

Serial dilution experiments were conducted using a single positive SARS-CoV-2 nasopharyngeal swab specimen. The viral transport media (VTM) was diluted serially between 10 to 1,000,000-fold. The dilutions were tested in triplicate to determine the results of LAMP and two reference RT-PCR methods (Envelope [E] gene and Nucleocapsid [N] 2 gene). Both RT-PCR and the RT-LAMP (S gene) amplified the target up to a dilution of 100,000 in a head-to-head comparison, suggesting equal LOD (data not shown). The same experiment was conducted comparing E gene RT-PCR to RT-LAMP (dual-target S and RdRP gene) as well as a direct RT-LAMP (dual-target S and RdRP gene) without formal extraction (boil method) on a separate specimen. These data are shown in Figure 5 and Table 4. Reproducible amplification occurred at 1000-fold dilution for RT-PCR (E gene) and RT-LAMP. Direct-LAMP amplified reproducibly at a 100-fold dilution for this sample.

**Figure 5:**
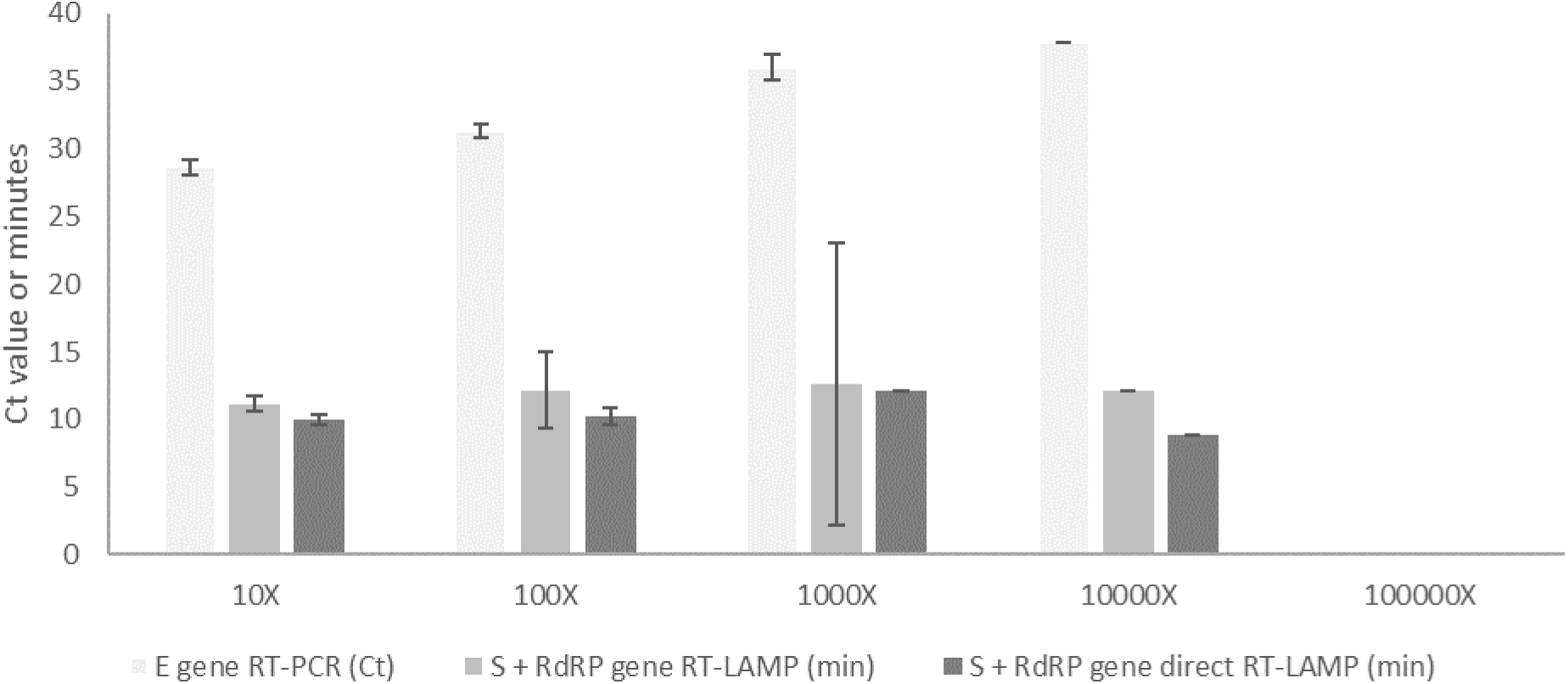
Serial dilution studies of a clinical sample (NP swab in VTM, E gene Ct 25.6) comparing the limit of detection of Envelope (E) gene RT-PCR (n=6), S gene + RdRP gene RT-LAMP (n=9) and S gene + RdRP gene direct RT-LAMP (n=9). Cycle threshold (Ct) value (RT-PCR) or time in minutes (LAMP) shown on y-axis. No virus was detected at a dilution of 1 × 10^5^ from the original neat sample. Standard deviation of the mean indicated in error bars if more than two values existed.

**Table 4:** Dual-target RT-LAMP (S and RdRP gene) assay results on a NP VTM sample (E gene Ct 25.6) containing SARS-Co-V2 compared to RT-PCR after kit-based RNA extraction and directly from a sample. Positive tested replicates/total number of replicates from repeated triplicate experiments is shown. Dilutions are based on 10-fold serial dilutions from a neat NP VTM sample.

| Sample dilution factor | E gene<br>RNA extract<br>RT-PCR | Dual RT-LAMP<br>RNA extract | Direct dual RT-LAMP<br>3 min/ 95°C |
| --- | --- | --- | --- |
| 10 | 6/6 | 9/9 | 9/9 |
| 100 | 6/6 | 9/9 | 9/9 |
| 1000 | 6/6 | 5/9 | 1/9 |
| 10000 | 1/6 | 1/9 | 0/9 |
| 100000 | 0/6 | 0/9 | 0/9 |

### Clinical validation of SARS-CoV-2 LAMP using nasopharyngeal swab samples

A sample set of nasopharyngeal swabs (n=108) from COVID-positive, COVID-negative, together with samples for other respiratory viruses were used to validate the S gene LAMP primer set (Table 5). Given no gold standard exists, percent positive agreement (PPA) and negative percent agreement (NPA) were calculated. Reference methods included RT-PCR (E gene and N2 gene) methods employed by reference laboratories. Using the RT-PCR (E gene) as a reference standard, there was 41/42 (97.62% (95% CI 87.43 - 99.94)) PPA and 77/78 (98.72% (95% CI 93.06 - 99.97)) NPA for the S gene LAMP. No cross-reactivity was observed for human coronaviruses (HCoV) OC43, 229E, NL63, and HKU1 or influenza virus A (H1N1) pdm09. Resolution of discrepant results (Table 6) revealed that one falsely positive LAMP result was positive based on the original RT-PCR result reported at the time of clinical testing and therefore represented a true positive that may be due to sample decay in storage. The second falsely negative sample was positive by all methods including the RdRP LAMP primer set and was deemed a false negative in the final analysis. In order to eliminate the concern for S gene false negatives, dual-target S and RdRP LAMP was performed to increase clinical sensitivity. This second clinical sample set intentionally included low positive (high Ct value) specimens. Table 7 demonstrates PPA of 44/48 (91.67% (95% CI 80.02 - 97.68)) and NPA 72/72 (100.00% (95% CI 95.01 - 100.00)) for the dual LAMP compared to the RT-PCR E gene reference method. Discrepant analysis (Table 8) revealed that all four falsely negative samples by LAMP were in fact positive by the CDC N2 RT-PCR, confirming the false negative status. The four discrepant specimens were repeated with single-target S gene LAMP and were also negative.

**Table 5:** Validation of RT-LAMP (S gene) compared to RT-PCR (E gene) for clinical samples (NP swabs), contrived samples, and negative control samples. PPA - positive percent agreement; NPA – negative percent agreement

| RT-LAMP (S gene) | RT-PCR (E gene) |  | Total |
| --- | --- | --- | --- |
|  | Positive | Negative |  |
| Positive | 41 | 1 | 42 |
| Negative | 1 | 77 | 78 |
| Total | 42 | 78 | 120 |
| PPA | 97.67% (95% CI 87.43 - 99.94) |  |  |
| NPA | 98.72% (95% CI 93.06 - 99.97) |  |  |

**Table 6:**
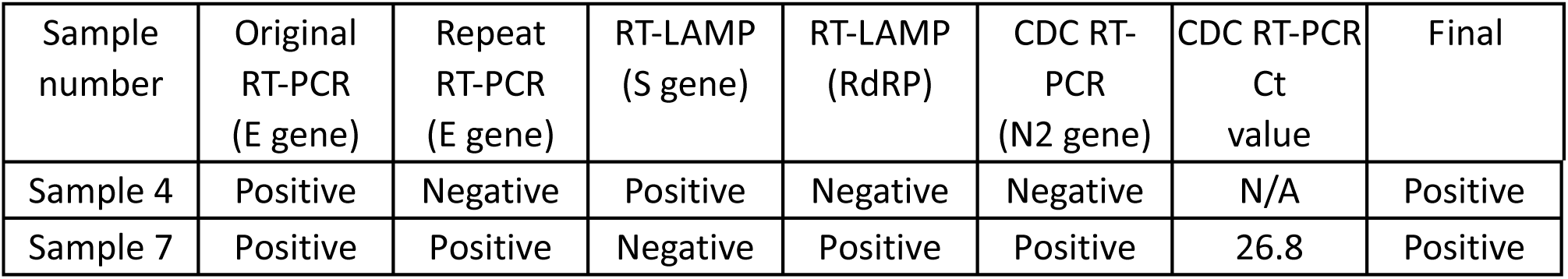
Discrepant analysis for single-target (S gene) RT-LAMP from the clinical validation data set.

**Table 7:** Validation of dual-target RT-LAMP (S gene and RdRP genes) using a validation set of clinical samples. PPA - positive percent agreement; NPA - negative percent agreement

| RT-LAMP (S + RdRP) | RT-PCR (E gene) |  | Total |
| --- | --- | --- | --- |
|  | Positive | Negative |  |
| Positive | 44 | 0 | 44 |
| Negative | 4 | 72 | 76 |
| Total | 48 | 72 | 120 |
| PPA | 91.67% (95% CI 80.02 - 97.68) |  |  |
| NPA | 100.00% (95% CI 95.01 - 100.00) |  |  |

**Table 8:** Discrepant analysis for dual-target (S + RdRP gene) RT-LAMP for samples with high Ct values from the clinical validation data

| Sample number | Original RT-PCR (E gene) | Repeat RT-PCR (E gene) | RT-LAMP (S + RdRP gene) | RT-LAMP (S gene) | CDC RT-PCR (N2 gene) | CDC RT-PCR Ct value | Final |
| --- | --- | --- | --- | --- | --- | --- | --- |
| Sample A7 | Positive | Positive | Negative | Negative | Positive | 38.36 | Positive |
| Sample A8 | Positive | Positive | Negative | Negative | Positive | 36.19 | Positive |
| Sample B7 | Positive | Positive | Negative | Negative | Positive | 36.41 | Positive |
| Sample D5 | Positive | Positive | Negative | Negative | Positive | 35.39 | Positive |

## Discussion

The global pandemic with SARS-CoV-2 has resulted in the need for diagnostic test development at a scale never seen before. Rapid deployment of validated laboratory-developed diagnostic tests or commercial tests was essential to the containment of the virus as it allows for self-quarantine measures to be imposed in a strategic fashion before widespread community transmission occurs (6, 7). Diagnostic tests have to be analytically sensitive in order to not to miss any cases in the acute phase of viremia (11). As such, NAATs serve this purpose. In particular, RT-PCR has been employed as the primary diagnostic counter-measure (20). However, reagent supply chains for key items are under immense pressure. Local solutions to reagent sources have become paramount because barriers to trade of these selected items have been a concern.

We noted that a one false negative occurred with single-target LAMP (S gene) for a lower Ct value sample that may be due to genetic polymorphism at key residues where LAMP primers bind. This issue was overcome by using two targets (S and RdRP genes) simultaneously. The single and dua-target RT-LAMP test for SARS-CoV-2 has comparable analytical sensitivity and achieved excellent agreement with the reference method. We noted that at high Ct value (>35), and presumably low-level infection, E gene RT-PCR identified specimens that LAMP did not. This suggests the LOD of the RT-PCR used in this study may be superior to LAMP for these low positives. However, we note that samples with E gene RT-PCR Ct values greater than 35 are relatively rare (~1%) at our reference laboratory (our unpublished observations). Increasing LAMP reaction times may reduce false negatives, but also lead to spurious amplification. The clinical relevance of these low positives is still not well understood and could represent early infection during the incubation period, late infection after the initial viral peak, or asymptomatic carriage. The transmissibility of these low positives to others is also not well understood.

Moreover, if we assume an overall prevalence of 5%, the negative predictive value of LAMP is 99.56% (95% CI 98.89 - 99.83) and positive predictive value is 100%. These values clearly support the use of LAMP as an alternative NAAT.

LAMP does not rely on the same reagents as RT-PCR and thus alleviates pressure on key supply chain items. The LAMP method is amenable to high throughput testing in either 96-well or 384-well. The assay is also able to detect SARS-CoV-2 in VTM without the need for a kit-based RNA extraction method relying on commercial reagents. However, we noted a 2-log drop in analytical sensitivity when direct LAMP was performed following this heat step. The drop in analytical sensitivity will only affect low-level viral load specimens. This is still within a good range compared to RT-PCR using RNA isolation. We believe that addition of a buffer to stabilize the RNA-enzyme complex in the LAMP reaction may further enhance this extraction-free approach. This needs to be tested. The heat step at 95°C for 3 minutes should significantly inactivate the virus permitting safe operation of the assay in a Class II biosafety cabinet (21). Taken together, these data support the use of LAMP chemistry as an alternate method for laboratory developed NAATs.

Our studies with a LAMP enzyme called GspSSD2 also provided encouraging results. These data demonstrated that lyophilized GspSSD2 and reagents are able to amplify SARS-CoV-2 directly from a specimen without a kit-based RNA extraction. Additionally, visual detection with a simple blue LED light is able to discriminate positive from negative. These features are particularly useful for resource-limited settings without sophisticated laboratory infrastructure or where the cost of or access to kit-based reagents and equipment are prohibitive. Further studies are required to clinically validate this low-cost approach.

Limitations of the study include not testing other sample types such as alternate swabs, nasal washes, oropharyngeal samples, sputum, or stool. This work is ongoing with a special emphasis on swab-free testing. Also formal SARS-CoV-2 viral titers were not calculated in the limit of detection studies. Nevertheless, LAMP presents a much needed alternative approach to SARS-CoV-2 diagnostic testing that is available for deployment immediately in a LDT format as it relies on other key reagents that do not cannibalize RT-PCR reagents. Ultimately, the aim is to port LAMP chemistry on a stand-alone microfluidic device POCT to be deployed in the community, either at ports of entry, homes, pharmacies, or resource-limited settings.

## Data Availability

Available upon reasonable request from the corresponding author.

## Acknowledgments

Dr. Ranmalee Amarasekara for expert technical assistance, Daniel Castaneda Mogollon for performing bioinformatics analysis, and Omar Abdullah for research analytical support.

